# Prehospital Stroke Scales Outperform National Institutes of Health Stroke Scale in Predicting Large Vessel Occlusion in a Large Academic Telestroke Network

**DOI:** 10.1101/2023.04.19.23288844

**Authors:** Stephen W. English, Nikita Chhabra, Abigail E. Hanus, Rida Basharath, Monet Miller, Richard J. Butterfield, Nan Zhang, Bart M. Demaerschalk

## Abstract

**Background:** With growing emphasis on prehospital identification of large vessel occlusion (LVO), some experts have advocated for prehospital involvement of vascular neurologists. Prehospital telestroke may improve triage and in-hospital treatment, but the accuracy of prehospital LVO scales in telestroke has not been investigated.

**Methods:** We performed a retrospective study of telestroke consultations in a large academic telestroke network from 2019 to 2020. We assessed performance of 7 LVO scales using the NIHSS score at presentation (RACE, C-STAT, FAST-ED, 3I-SS, PASS, VAN, and G-FAST). We performed two analyses using different LVO definitions: (1) anterior LVO including occlusion of the internal carotid (ICA), middle cerebral (M1 or M2), or anterior cerebral (A1 or A2) arteries; and (2) any LVO including occlusion sites above plus basilar artery or posterior cerebral artery (P1 or P2). Diagnostic performance was assessed via sensitivity, specificity, negative predictive value, positive predictive value, positive likelihood ratio, negative likelihood ratio, and accuracy using established thresholds of each scale. These results were compared to NIHSS at thresholds of 6, 8, and 10. Area under curve (AUC) was calculated using c-statistics by treating scales as continuous variables.

**Results:** A total of 625 patients were included; 111 (17.8%) patients had an anterior LVO, 118 (18.9%) patients had any LVO, and 182 (29.1%) patients had stroke mimic diagnosis. Mean age (SD) was 67.9 (15.9), 48.3% were female, and 93.4% were white. Mean NIHSS (SD) was 14.9 (8.4) for patients with anterior LVO, 4.7 (5.0) for patients with non-LVO ischemic stroke, and 4.4 (5.8) for stroke mimic (p<0.001). Compared to the NIHSS, FAST-ED and RACE scales demonstrated higher accuracy and AUC for LVO detection.

**Conclusions:** Both the FAST-ED and RACE scales outperformed the NIHSS for LVO detection in patients evaluated by telestroke. These scales may be valid alternatives to the NIHSS examination in this setting.

## Introduction

Endovascular therapy (EVT) is highly effective in acute ischemic stroke (AIS) due to large vessel occlusion (LVO),^1-6^ but this treatment is only offered in comprehensive stroke centers (CSCs) or thrombectomy-capable stroke centers (TSCs). Emergency Medical Services personnel (EMS)- administered LVO detection scales and prehospital bypass protocols have been developed to improve prehospital LVO detection and triage. The most common prehospital LVO scales include the Los Angeles Motor Scale (LAMS),^7^ Rapid Arterial oCclusion Evaluation scale (RACE),^8^ Cincinnati Prehospital Stroke Scale (C-STAT),^9^ Field Assessment Stroke Triage for Emergency Destination scale (FAST-ED),^10^ 3-Item Stroke Scale (3I-SS),^11^ Vision-Aphasia-Neglect scale (VAN),^12^ Gaze-Face-Arm-Speech-Time scale (G-FAST),^13^ and Prehospital Acute Stroke Severity Scale (PASS).^14^ Despite widespread implementation, these scales have limited accuracy^15,16^ and do not incorporate other clinical factors that impact treatment decisions. Therefore, some experts have advocated for greater involvement of vascular neurologists in the prehospital setting as seen with the introduction of mobile stroke units (MSUs)^17,18^ and prehospital telemedicine^19-22^ to accelerate triage and treatment decisions.

With the increasing utilization of prehospital telestroke, there is growing demand for rapid and accurate assessment tools to guide neurologists in the triage of patients with AIS. The common prehospital LVO scales are abbreviated or partial versions of the National Institutes of Health Stroke Scale (NIHSS) focusing on the highest yield components, and while no direct comparisons have been made in the literature to compare the timeliness of each scale, it can be assumed that these scales are completed faster than a full NIHSS. Despite this advantage, the accuracy of these prehospital LVO scales have not been formally evaluated in telestroke practice. Therefore, we sought to determine the accuracy of common LVO scales in a large academic telestroke practice.

## Methods

### Setting

The Mayo Clinic Telestroke program was established in 2007 to provide comprehensive evidence-based care to patients in a hub-and-spoke paradigm around their comprehensive stroke centers in Minnesota, Florida, and Arizona. The program has grown to include 27 sites in 2021, of which 17 are Mayo Clinic Health System (MCHS) sites in rural Minnesota and Wisconsin that serve as the spoke sites for the CSC site at Mayo Clinic in Rochester, Minnesota. All neurologists performing telestroke evaluations are board-certified academic vascular or critical care neurologists at Mayo Clinic. Telestroke activation criteria includes all patients with acute focal neurologic deficits within 24 hours from last known well.

Other details concern the structure, personnel, staffing, workflow, and operations have been previously reported.^23,24^

### Study Population

We performed a retrospective chart review of consecutive video telestroke consultations performed at MCHS sites from January 1, 2019 to December 31, 2020. Patients at network sites external to Mayo Clinic were excluded due to disparate electronic health records. Additional exclusion criteria included: age less than 18 years old at time of telestroke, no NIHSS documentation, no vessel imaging performed, no research authorization, or LVO deemed to be chronic or asymptomatic. Chart review was conducted (S.E., N.C., A.H., R.B., M.M.) to abstract the following: age on presentation, gender, race and ethnicity, vascular risk factors, presenting NIHSS score, initial blood pressure, initial blood glucose, imaging characteristics including type of vessel imaging, presence of LVO, site of occlusion, and whether intravenous thrombolysis (IVT) or EVT were recommended and received. Patients with hyperdense middle cerebral artery (MCA) who did not receive vascular imaging were included if the telestroke neurologist suspected an acute LVO. This study was reviewed and determined to be exempt by the Mayo Clinic Institutional Review Board.

### LVO Definition

We chose to complete 2 different analyses based on how LVO was defined. The first definition, referred to as the anterior circulation criteria, focused only on patients with anterior circulation LVO in line with prior publications^16,25^ and included occlusion of the internal carotid artery (ICA), middle cerebral artery (MCA) M1 or M2 branches, and anterior cerebral artery (ACA) A1 or A2 branches. The second definition attempted to capture any patient who may be considered for mechanical thrombectomy based on site of occlusion and severity of deficits in current clinical practice. This definition, referred to as the posterior circulation criteria, includes occlusion of the ICA, M1, M2, A1, A2, basilar artery, or posterior cerebral artery (PCA) P1 or P2 branches. Two neurologists (S.E. and N.C.) reviewed patient records of each LVO case to ensure that chronic incidental LVOs were excluded.

### LVO Recognition Scales

Prehospital LVO scales were calculated retrospectively based on the results of the NIHSS at time of telestroke encounter. Only scales that could be extrapolated from the presenting NIHSS were included in analysis (due to incorporation of grip strength, the LAMS score^7^ was not included). The following scales were evaluated: NIHSS, RACE, C-STAT, FAST-ED, 3I-SS, VAN, G-FAST, and PASS. Twenty-two patients had symmetric weakness on examination; therefore, the RACE score could not be applied in these cases. In cases there was asymmetric weakness, the side with more significant weakness was used to calculate scores.

### Statistical Analysis

Data was organized into continuous and categorical variables. Baseline demographic and presenting clinical information were categorized according to diagnosis and described as mean and standard deviation (SD) for continuous variables or as frequency and percentage for categorical variables. These data were subsequently grouped to compare based on diagnosis: anterior circulation LVO, non-anterior circulation ischemic stroke or TIA, or stroke mimic. Diagnostic performance was assessed by calculating sensitivity, specificity, negative predictive value (NPV), positive predictive value (PPV), accuracy, positive likelihood ratio, and negative likelihood ratio using the established thresholds of each scale. These results were compared to NIHSS at thresholds of 6, 8, and 10. Area under curve (AUC) was calculated using c-statistics by treating scales as continuous variables. All analyses were conducted with R version 4.1.2 (R Foundation for Statistical Computing, Vienna, Austria). P-value of 0.05 was chosen as the cut-off criterion for statistical significance.

## Results

There were 1055 video telestroke encounters that met initial inclusion criteria during our study period. Of these, 397 did not have vascular imaging and 33 did not have NIHSS documentation in the telestroke encounter. A total of 625 patients were included in the analysis, of which 111 patients (17.8%) were diagnosed with anterior circulation LVO, 118 (18.9%) patients were diagnosed with any LVO, and 182 patients (29.1%) were diagnosis as a stroke mimic. Baseline characteristics and initial presenting data are reported in Table 1. Mean age (SD) was 67.9 (15.9), 302 (48.3%) patients were female, and 582 (93.4%) patients were white. Patients with anterior LVO were more likely to have atrial fibrillation (32.4% vs 18.7% vs 11.0%), p<0.001), less likely to use tobacco (15.5% vs 19.9% vs 27.1%), have a higher NIHSS (14.9 vs 4.7 vs 4.4,p<0.001), and had a lower presenting systolic blood pressure (149.3 mmHg vs 158.5 mmHg vs. 152.3 mmHg, p =0.002) compared patients with non-LVO ischemic stroke or stroke mimic diagnosis, respectively. Imaging source for LVO determination was CT angiography in 571 (91.4%) patients, MR angiography in 34 (5.4%) patients, and CT scan with hyperdense MCA sign in 20 (3.2%) patients. There were 123 vessel occlusions identified in 118 patients; 5 patients had multiple vascular occlusions in different territories. The location of all vessel occlusions in our cohort is reported in Table 3. Immediate vessel imaging at time of presentation was obtained in 413 (67.2%) cases, whereas CT perfusion was only available in 30 (4.8%) patients. Telestroke neurologists recommended IVT in 162 patients (25.9%) and transfer for possible mechanical thrombectomy in 128 cases (20.5%). Ultimately, IVT was administered in 154 patients (24.6%) and thrombectomy was ultimately performed in 55 patients (8.8%).

**Table 1:** Baseline Characteristics.

|  | **LVO Ischemic Stroke**<br>**(n=111)** | **Non-LVO**<br>**Ischemic Stroke**<br>**(n=332)** | **Stroke Mimic**<br>**(n=182)** | **Total**<br>**(n=625)** | **p-**<br>**value** |
| --- | --- | --- | --- | --- | --- |
| Age | 71.9 (15.6) | 70.3 (14.1) | 61.0 (17.0) | 67.9(15.9) | <0.001 |
| Female | 55 (49.5) | 143 (43.1) | 104 (57.1) | 302 (48.3) | 0.009 |
| White | 104 (93.7) | 313 (94.3) | 167 (91.8) | 584 (93.4) | 0.196 |
| Risk Factors (n, %) |  |  |  |  |  |
| Hypertension | 81 (73.0) | 262 (78.9) | 113 (62.1) | 456 (73.0) | <0.001 |
| Hyperlipidemia | 78 (70.3) | 234 (70.5) | 84 (46.2) | 396 (63.4) | <0.001 |
| Diabetes Mellitus | 32 (28.8) | 100 (30.1) | 53 (29.1) | 185 (29.6) | 0.954 |
| Coronary Artery Disease | 34 (30.6) | 94 (28.3) | 34 (18.7) | 162 (25.9) | 0.027 |
| Atrial Fibrillation | 36 (32.4) | 62 (18.7) | 20 (11.0) | 118 (18.9) | <0.001 |
| Tobacco Use | 17 (15.5) | 66 (19.9) | 49 (27.1) | 132 (21.2) | 0.065 |
| Prior ischemic stroke | 20 (18.0) | 82 (24.7) | 27 (14.8) | 129 (20.6) | 0.023 |
| Presentation (mean, [SD]) |  |  |  |  |  |
| NIHSS | 14.9 (8.4) | 4.7 (5.0) | 4.4 (5.8) | 6.4 (7.1) | <0.001 |
| Systolic Blood Pressure | 149.3 (24.0) | 158.5 (27.3) | 152.3 (26.2) | 155.1 (26.7) | 0.002 |
| Blood glucose | 145.5 (85.2) | 135.8 (60.5) | 140.1 (95.2) | 138.8 (76.6) | 0.492 |
| *LVO defined using the anterior circulation criteria |  |  |  |  |  |

**Table 2:** Site of Large Vessel Occlusion.

| **Location** | **n = 123 (%)** |
| --- | --- |
| Internal Carotid Artery |  |
| Left | 8 (6.5) |
| Right | 8 (6.5) |
| Middle Cerebral Artery |  |
| Left M1 | 26 (21.1) |
| Right M1 | 24 (19.5) |
| Left M2 | 13 (10.6) |
| Right M2 | 20 (16.3) |
| Tandem |  |
| Left | 6 (4.9) |
| Right | 3 (2.4) |
| Anterior Cerebral Artery |  |
| Left A1/A2 | 2 (1.6) |
| Right A1/A2 | 1 (0.8) |
| Basilar Artery | 3 (2.4) |
| Posterior Cerebral Artery |  |
| Left P1/P2 | 5 (4.1) |
| Right P1/P2 | 4 (3.3) |

**Table 3:**
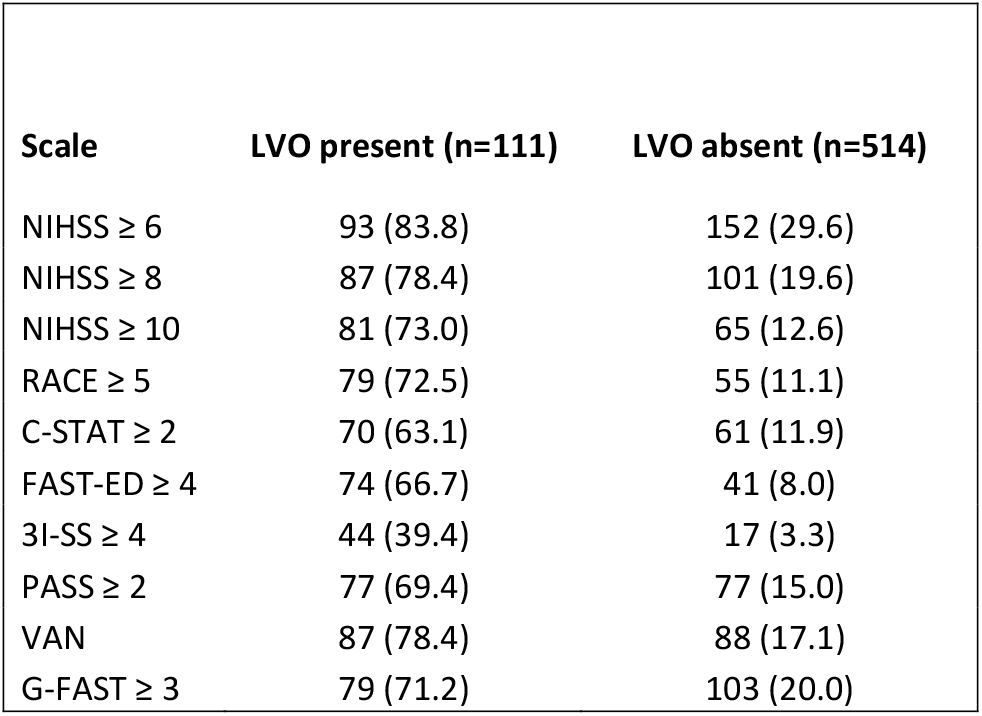
Prehospital LVO scale scores based on anterior circulation LVO status (n, %)

| **Scale** | **LVO present (n=111)** | **LVO absent (n=514)** |
| --- | --- | --- |
| NIHSS $\geq 6$ | 93 (83.8) | 152 (29.6) |
| NIHSS $\geq 8$ | 87 (78.4) | 101 (19.6) |
| NIHSS $\geq 10$ | 81 (73.0) | 65 (12.6) |
| RACE $\geq 5$ | 79 (72.5) | 55 (11.1) |
| C-STAT $\geq 2$ | 70 (63.1) | 61 (11.9) |
| FAST-ED $\geq 4$ | 74 (66.7) | 41 (8.0) |
| 3I-SS $\geq 4$ | 44 (39.4) | 17 (3.3) |
| PASS $\geq 2$ | 77 (69.4) | 77 (15.0) |
| VAN | 87 (78.4) | 88 (17.1) |
| G-FAST $\geq 3$ | 79 (71.2) | 103 (20.0) |

Prehospital LVO scale scores are summarized as dichotomized groups according to presence or absence of anterior circulation LVO (Table 3). The diagnostic performance of prehospital LVO scales is outlined based on the anterior circulation criteria (Table 4) and posterior circulation criteria (Table 5). NIHSS ≥ 6 was the most sensitive (83.8% and 82.2%) and had strongest NPV (95.3% and 94.5%), whereas 3I-SS ≥ 4 was the most specific (96.7% and 96.6%) and had strongest PPV (72.1% and 72.1%), using the anterior and posterior circulation LVO criteria, respectively. Of the prehospital LVO scales evaluated (excluding NIHSS), VAN was the most sensitive in both the anterior circulation (78.4%) and posterior circulation criteria (76.3%). Three prehospital LVO scales demonstrated higher accuracy than NIHSS using both criteria: RACE ≥ 5, FAST-ED ≥ 4, and 3I-SS ≥ 4. However, only two scales demonstrated higher AUC than NIHSS (85.6% and 85.0%): RACE (86.1% and 85.0%) and FAST-ED (87.4% and 86.2%) for anterior and posterior circulation definitions, respectively. Based on the AUC results, receiver operator characteristic (ROC) curves were generated based on both the anterior circulation (Figure 1) and posterior circulation criteria (Figure 2).

**Table 4.** Diagnostic Performance of Prehospital LVO Scales using Anterior Circulation Criteria.

| Scale | Sensitivity<br>(%) | Specificity<br>(%) | PPV<br>(%) | NPV<br>(%) | Accuracy<br>(%) | AUC<br>(%) | LR+ | LR- |
| --- | --- | --- | --- | --- | --- | --- | --- | --- |
| NIHSS $\geq 6$ | 83.8 | 70.4 | 38.0 | 95.3 | 72.8 | 85.6 | 2.83 | 0.23 |
| NIHSS $\geq 8$ | 78.4 | 80.4 | 46.3 | 94.5 | 80.0 | 85.6 | 3.99 | 0.27 |
| NIHSS $\geq 10$ | 73.0 | 87.4 | 55.5 | 93.7 | 84.8 | 85.6 | 5.77 | 0.31 |
| RACE $\geq 5$ | 72.5 | 88.9 | 59.0 | 93.6 | 85.9 | 86.1 | 6.51 | 0.31 |
| C-STAT $\geq 2$ | 63.1 | 88.1 | 53.4 | 91.7 | 83.7 | 81.4 | 5.31 | 0.42 |
| FAST-ED $\geq 4$ | 66.7 | 92.0 | 64.3 | 92.7 | 87.5 | 87.4 | 8.36 | 0.36 |
| 3I-SS $\geq 4$ | 39.6 | 96.7 | 72.1 | 88.1 | 86.6 | 80.9 | 11.99 | 0.62 |
| PASS $\geq 2$ | 69.4 | 85.0 | 50.0 | 92.8 | 82.2 | 82.3 | 4.63 | 0.36 |
| VAN | 78.4 | 82.9 | 49.7 | 94.7 | 82.1 | N/A | 4.58 | 0.26 |
| G-FAST $\geq 3$ | 71.2 | 80.0 | 43.4 | 92.8 | 78.4 | 83.9 | 3.55 | 0.36 |

\*PPV, positive predictive value; NPV, negative predictive value; LR+, positive likelihood ratio; LR-, negative likelihood ratio.

**Table 5.**
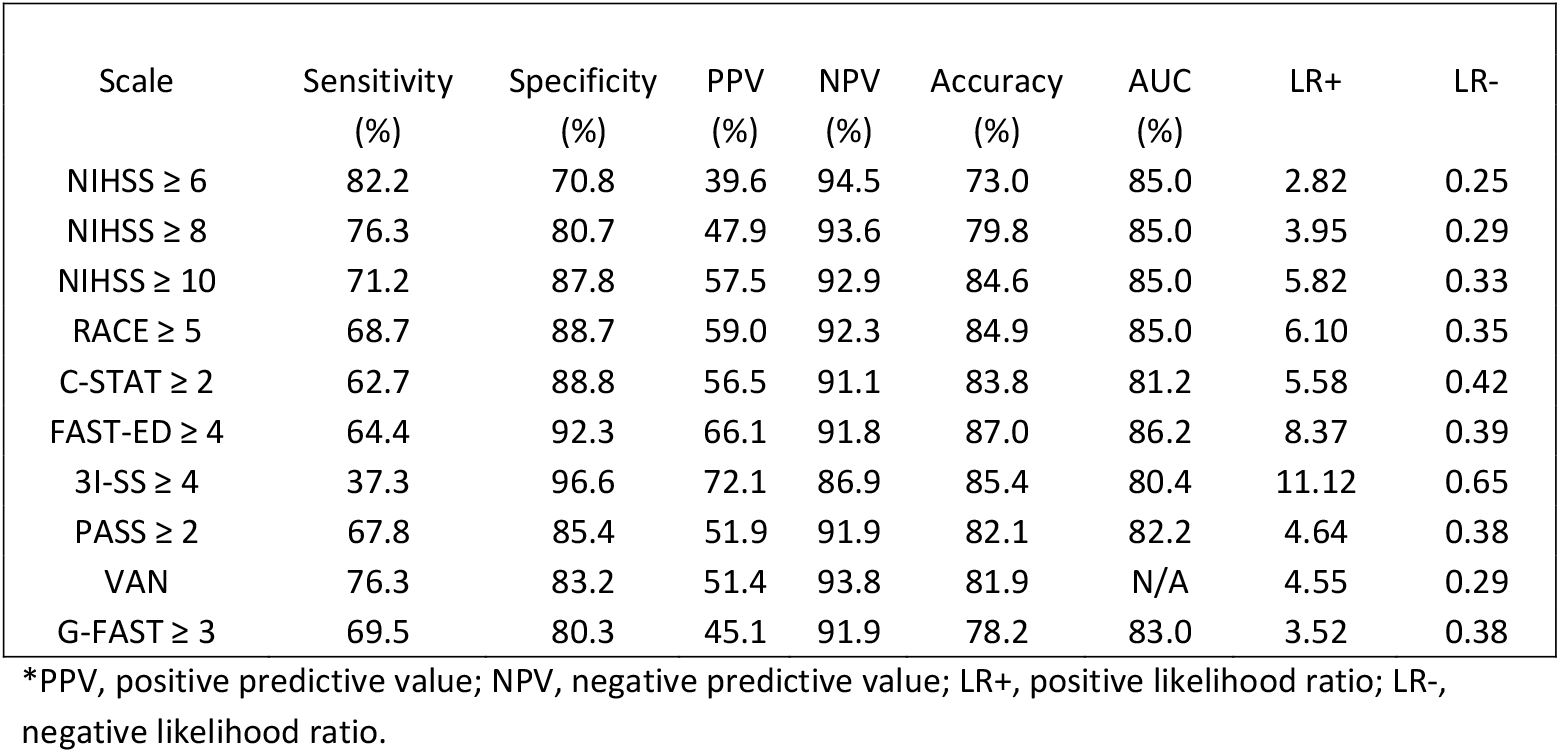
Diagnostic Performance of Prehospital LVO Scales using Posterior Circulation Criteria.

**Figure 1.**
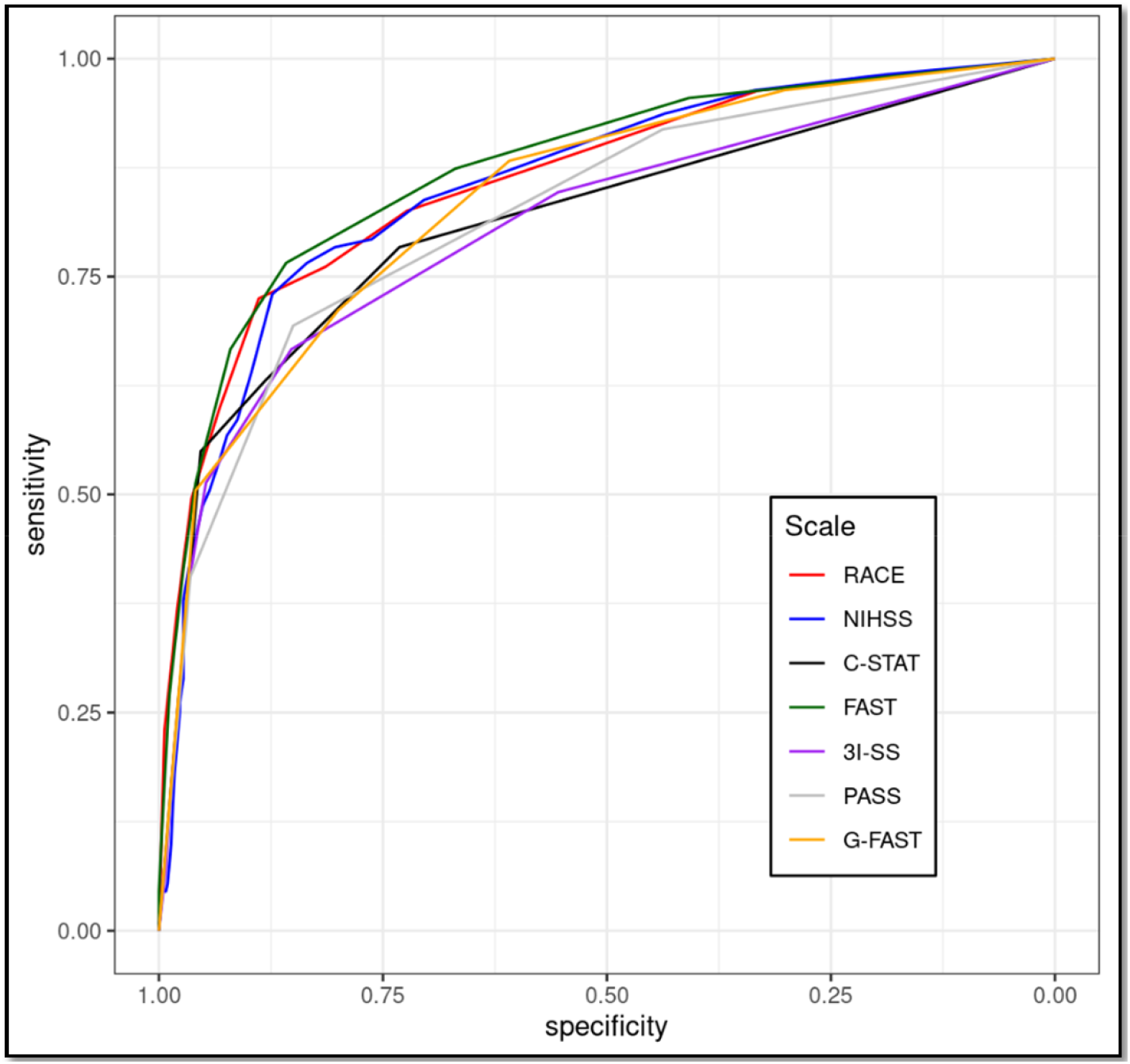
ROC Curves for Prehospital LVO Scales using Anterior Circulation Criteria.

**Figure 2.**
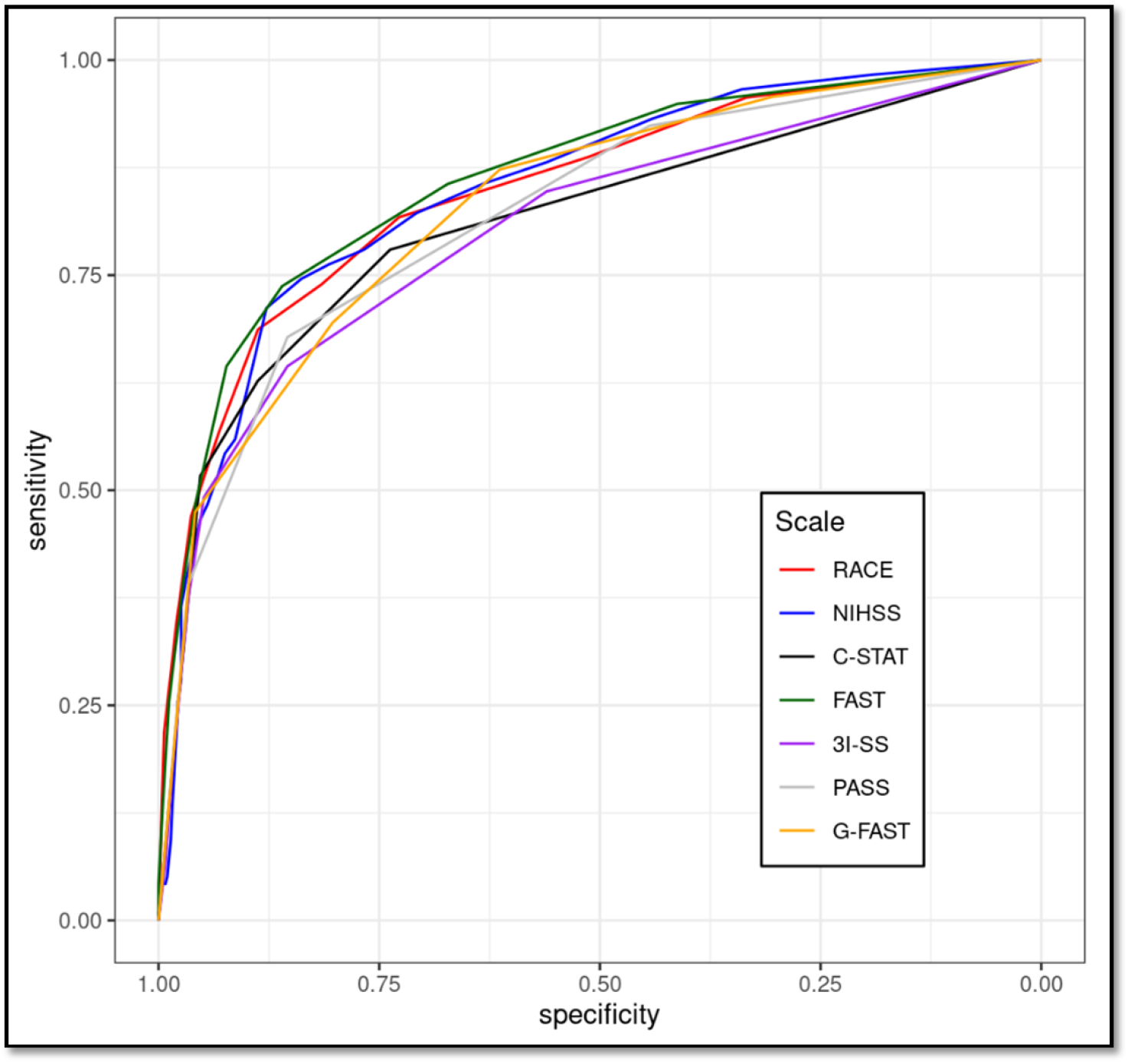
ROC Curves for Prehospital LVO Scales using Posterior Circulation Criteria.

## Discussion

To our knowledge, this is the first study to evaluate the accuracy of prehospital LVO scales in a large academic telestroke network. Our study highlights two prehospital LVO scales, the FAST-ED and RACE scales, that outperform the NIHSS in recognizing LVO when administered via telestroke by a vascular neurologist. These results reinforce findings that have been previously reported elsewhere in the literature. One systematic review of prehospital LVO scale performance identified the RACE (AUC 0.82) and FAST-ED scales (AUC 0.83) as the only two LVO detection scales to outperform the NIHSS (AUC 0.81) when administered by EMS personnel in the ambulance.^25^ A similar study by Anadani et al.^26^ sought to evaluate the accuracy of LVO scales in telestroke and again identified that the FAST-ED (AUC 0.81) and RACE (AUC 0.82) scales were the only two LVO scales to outperform the NIHSS (AUC 0.77).

However, this study only included patients who were already transferred to a TSC for suspected LVO, resulting in a cohort of patients with higher percentage of LVO (48.5%) and a much lower percentage of stroke mimics (6.2%). This population is not representative of patients that would routinely present via ambulance to the emergency department as a possible stroke, so we believe that our study helps to further support the role of these scales as valuable tools in prehospital stroke triage.

Prehospital LVO detection scales have become the most widely used triage tools for AIS due to their low-cost and ease-of-use, but reports of their accuracy in the real-world setting are mixed.(20) Some of the more subtle findings associated with LVO, including visual field deficits and agnosia, have proven difficult for EMS personnel to interpret, thereby limiting the applicability of some of the highest performing scales.(21) As such, several centers are evaluating the role of prehospital telemedicine performed by a vascular neurologist to improve triage and reduce in-hospital treatment delays.^19,20,26^ Our study reveals that even the top-performing scales administered by a vascular neurologist in the telestroke setting have significant room for improvement. All the scales have a high NPV greater than 90%, but this is largely driven by the low rates of LVO in the population. Despite using the more inclusive posterior circulation criteria for LVO, the PPV of the highest performing scales RACE and FAST-ED have a PPV of 59% and 66.1%, respectively. This suggests that a vascular neurologist led telemedicine prehospital triage system would still result in more than a third of patients without LVO meeting bypass criteria. This can cause downstream strain on a tertiary care facility and potentially delay IVT in eligible patients. MSUs with on-board vascular imaging can help mitigate this false positive rate. Several studies have demonstrated the role of CT angiography in a MSU can lead to 100% accurate triage rates due to the angiographic confirmation of a proximal LVO prior to triage decision. ^27,28^ Due to cost and staffing limitations, the scalability and integration of these MSUs into existing prehospital EMS infrastructure has been called into question.^22^

One question to consider is how much value a vascular neurologist assessment adds compared to EMS personnel performance? By comparing the accuracy of overlapping scales from one large, prospective study by Nguyen et al.^16^ the accuracy ranged from 79%-88% in their study compared to 78%-88% in our study. While we would anticipate that vascular neurologists should outperform EMS personnel, these scores have high interrater reliability, the EMS personnel in the study received more in-depth training to participate in the study than in real-world settings, and the EMS personnel were able to perform the testing in a live environment rather than telemedicine. Another study took a more real-world cohort without any specialized EMS training to evaluate the accuracy of these scales;^25^ here, we see differences in the AUC with ranges in performance from 0.75-0.83 in the EMS group versus 0.80-0.87 in our study. Although these results suggest an overall improvement in accuracy with telemedicine vascular neurologist assessments, we argue that the appeal of prehospital telestroke extends beyond accuracy of these LVO scales. A trained vascular neurologist will be more capable of determining the presence of a posterior circulation LVO (included scales are weighted to capture anterior LVO) that benefit from bypass, recognizing subtle cortical signs such as agnosia, identifying clues that a presentation is more suggestive of a stroke mimic (e.g. post-ictal Todd’s paralysis) or alternative neurologic emergency (e.g. status epilepticus), and capturing critical information that will inform and expedite both prehospital triage and in-hospital care such as oral anticoagulation status, last known well time, family contact information, and baseline functional status. Not only do these aspects improve hospital triage and resource allocation, but it also allows for faster administration of reperfusion therapies due to the ability to shift components of in-hospital care to the prehospital setting.^20,22^

Beyond the prehospital setting, there may be other opportunities to leverage these scales to improve LVO recognition. Despite the widespread availability of CT angiography in primary stroke centers and Acute Stroke Ready hospitals in the United States, there are still many free-standing Emergency Departments (EDs) and small rural community EDs that do not have CT angiography available, and patients need to transfer out if there is any concern for acute ischemic stroke. Leveraging these scales with quick telemedicine assessments by a trained vascular neurologist may be an effective triage tool to determine need for emergent transfer, particularly in rural areas where air or ground transportation is less readily available and resource allocation is an important consideration.

Part of the challenge in comparing performance of these scales is the differences in how we define LVO. Many of the studies strictly focus on anterior circulation LVO as the scales are designed to assess for deficits attributable to anterior circulation ischemia.^8,25,26^ Although less common than anterior circulation LVO, it is reasonable to argue that recognition of acute basilar or proximal PCA occlusion may be just as important due to the morbidity and mortality associated with these LVO syndromes and there is growing evidence that EVT is the preferred treatment of choice for patients with these presentations.^29-31^ We chose to include both an anterior circulation and a more inclusive posterior circulation LVO definition for this reason, but due to the low numbers of posterior circulation LVOs in this cohort, it is difficult to extrapolate any meaningful conclusions from this analysis.

Our study has several limitations. This study is retrospective in nature and the prehospital LVO scales were calculated based on the NIHSS rather than determined prospectively in the telestroke encounter. Our patient population is predominantly white and does not reflect the racial and ethnic diversity in other areas of the United States, which may limit the generalizability of these findings. The presence of LVO may have been underrecognized due to several factors: lack of vascular imaging, limited utilization of CT perfusion, delays to obtaining vascular imaging, or administration of IV thrombolysis prior to obtaining vascular imaging.

This study identifies two prehospital LVO scales, the FAST-ED and RACE scale, that may outperform the NIHSS for detection of LVO in patients evaluated by telestroke. As prehospital telestroke utilization increases, the results of this study provide further insights into the critical aspects of the neurologic examination that can help to expedite triage without sacrificing accuracy. These scales may be valid alternatives to the NIHSS examination in this setting.

## Data Availability

The data that support the findings of this study are available from the corresponding author upon reasonable request.

## Acknowledgements

We would like to acknowledge Ms. Charisse Nord, Ms. Emily Pahl and all our telestroke providers (Dr. Kevin Barrett, Dr. Felix Chukwudelunzu, Dr. Bart Demaerschalk, Dr. Oana Dumitrascu, Dr. Stephen English, Dr. Kelly Flemming, Dr. William Freeman, Dr. Courtney Hrdlicka, Dr. Josephine Huang, Dr. Zafer Keser, Dr. James Klaas, Dr. Gyanendra Kumar, Dr. Michele Lin, Dr. Elizabeth Mauricio, Dr. Bayan Moustafa, Dr. Deena Nasr, Dr. Cumara O’Carroll, Dr. Alejandro Rabinstein, Dr. Lindsy Williams, Dr. Micah Yost) for their ongoing support and contributions to in telestroke network clinical operations, administration, and clinical consultations.

